# Distinct patterns of whole-body coding in human motor cortex and posterior parietal cortex

**DOI:** 10.1101/2025.08.01.25332521

**Authors:** Kelly Kadlec, Jorge Gamez, Tyson Aflalo, Charles Guan, Emily R. Rosario, Kelsie Pejsa, Ausaf Bari, Nader Pouratian, Richard Andersen

## Abstract

Traditional views of cortical motor areas hold that movements of different parts of the body, or effectors, are represented in distinct anatomical locations. The extent of overlap in effector representations has been a topic of debate for decades. We recorded from single neurons in “hand areas” of motor cortex (MC) and posterior parietal cortex (PPC) of tetraplegic humans while they attempted movements across the body.

We found a population response to every movement tested in both areas; however, the hand knob of MC more selectively activated for hand movements, whereas PPC did not emphasize any specific effector.

Single neurons in MC responded selectively to a single effector or to two effectors that were part of the same limb. In PPC, neurons responded to random combinations of effectors. These findings suggest a transition from highly effector-general representation in PPC to regional effector specificity in MC.

## Introduction

Even simple movements rely on the recruitment of multiple brain areas. Understanding the functional organization of these brain areas is a classic problem in motor control. The fundamental question of how different brain regions contribute to the control of different parts of the body has provided a framework to understand how information is arranged on the cortical surface more generally. In motor cortex (MC), results are summarized by the homunculus, an orderly representation of the body that is topographically arranged from head to toe ^1^. In posterior parietal cortex (PPC), results are summarized as distinct cortical regions responsible for the sensory-motor guidance of distinct effectors, such as the eyes in the lateral intraparietal area, the arm in parietal reach region, and the hand in anterior intraparietal area ^2–5^.

However, several recent results challenge this conventional wisdom. In human MC, recent functional magnetic resonance imaging (fMRI), stereoelectroencephalography (sEEG), and intracortical recordings all show a degree of overlap in effector representations ^6–9^. Although the original topographic organization did include some overlap between neighboring effectors (e.g. the fingers ^10^), the overlapping effectors in more recent findings are from head to toe. For example, single neuron recordings in humans from the hand knob of precentral gyrus found a compositional code for both the hand and the entire body^9^. Additionally, coding for both the ipsilateral and contralateral arm/hand are found to overlap in a single hemisphere (and even within single neurons) ^11–14^. In non-human primates (NHPs), long train intracortical microstimulation (LT-ICMS) studies have revealed complex movements, suggesting that MC is organized ethologically around behaviorally relevant effectors rather than the classical homunculus ^15–17^.

In PPC, recent fMRI studies in humans show overlapping representations for effectors for similar types of movements, suggesting a functional organization of movement ^18–21^. Previously, we recorded from human PPC and found that contralateral hand and shoulder movements were encoded orthogonally in single neurons whereas other variables were randomly mixed, an organization we referred to as partially mixed selectivity ^22, 23^. Additionally, LT-ICMS of PPC in NHPs can evoke multi-effector movements supporting the hypothesis of functional organization. ^24, 25^.

These recent findings of overlapping effector representations across various brain regions raise the question of whether different cortical areas encode movement and effectors in distinct or similar ways. Due to differences in tasks and participants in previous studies of whole body encoding in MC and PCC, it is difficult to directly compare these studies. In the current study, we had the unique opportunity to simultaneously human PPC and MC single neuron activity during attempted movements from head to foot in two participants. Examining these neural responses from each area in the same participant allows us to answer questions about how control of different effectors is organized in each area.

Despite targeting areas of both MC and PPC that have been classically associated with representing hand movements, we found neural responses to every part of the body; however, the pattern of this overlapping representation varied between the two brain regions. In MC, the population response for the wrist and thumb was two to three times stronger than other effectors. Most neurons were tuned to the contralateral hand, followed by the ipsilateral hand, with the fewest number of neurons being tuned to the proximal effectors. The population in PPC was found to represent the whole body as well, however, unlike in MC, we found equivalent tuning strength across effectors. There was no relationship observed between the effectors coded by each single neuron in PPC, and instead each neuron was tuned to a seemingly random combinations of effectors.

We also sought to understand whether this overlap has functionally useful characteristics, for example shared spatial information between effectors, which has been suggested to facilitate the transfer of learned motor skills from one limb to another. The extent to which information is shared is expected to vary between lower and higher cognitive areas, leading to a prediction that in MC spatial coding would be shared between specific effectors, such as the left and right hand, while in PPC arbitrary effectors would be able to share information ^22, 23^. Indeed, we find this to be the case. These results point to clearly distinct organization of effector representations in MC and PPC and allow for a greater understanding of the different functional roles of each area and how each area contributes to motor control.

## Results

As part of an ongoing clinical study, two participants were implanted with microelectrode Utah arrays. Participant N1 is tetraplegic (C3/C4) and was implanted with four 64-channel arrays in the left hemisphere: two in the hand knob of precentral gyrus (N1-MCL and N1-MCM), one in the left superior parietal lobule (SPL) of PPC (N1-PPC) and one in the supramarginal gyrus (see note in methods) (Figure 1a). Participant N2 is tetraplegic (C4/C5) and was implanted with two 96-channel arrays in the left hemisphere, one in the hand knob of precentral gyrus (N2-MC) and the other in SPL (N2-PPC, Figure 1b). Implant locations within each brain area were selected by finding the regions of peak activation in response to the participants performing motor imagery for hand and arm movements during functional magnetic resonance imaging.

**Figure 1.**
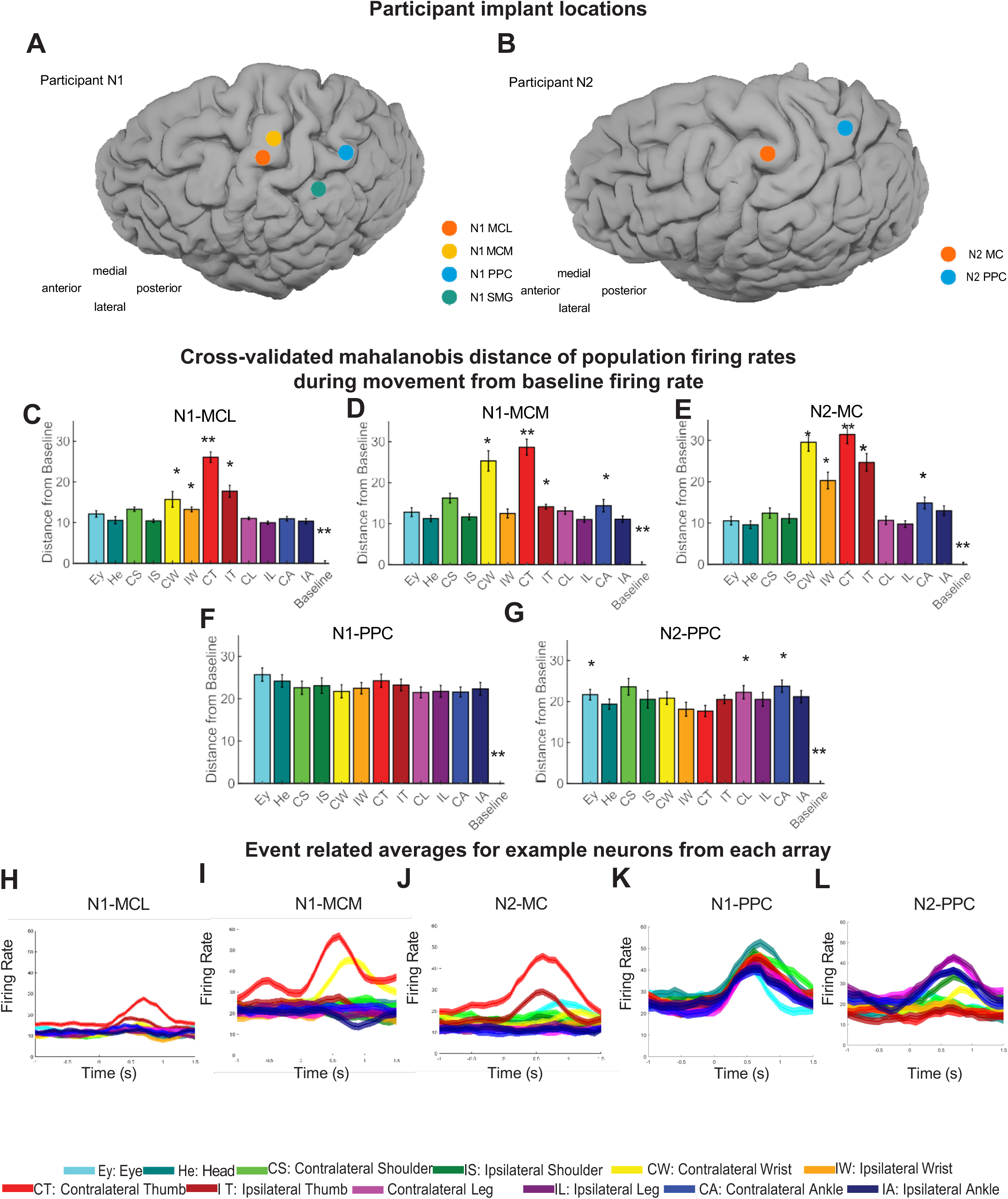
Fully body coding with distinct population responses in MC and PPC 1A: N1 Implant locations: N1 has two 64-channel Utah arrays located in the left hemisphere near the hand knob of precentral gyrus. The array that is located more medially is referred to as N1-MCM and the array that is more lateral is denoted N1-MCL. N1 has another 64-channel array in the left hemisphere located slightly medial and posterior to the junction of the postcentral and intraparietal sulcus, denoted N1-PPC. A final 64-channel array in the left supramarginal gyrus, N1-SMG, was not included in results because only a small number of neurons recorded from this array were found to be directionally tuned. 1B: N2 Implant locations: N2 has one 96-channel Utah array located in left hemisphere near the hand knob of precentral gyrus, denoted N2-MC. Additionally, N2 has one 96-channel array in the left hemisphere that is located slightly medial and posterior to the junction of the postcentral and intraparietal sulcus, referred to as N2-PPC. 1C-G: Cross-validated mahalanobis distance of population firing rates sampled from a 500ms window during movement execution and firing rates sampled from 500ms window during rest. Error bars show standard error of the mean. Significance was tested using a permutation test. Two stars indicate that the distance from baseline for that condition was significantly different from the distance from baseline for all other conditions. One star indicates that the distance from baseline for that condition was significantly different from that for select other conditions listed below. p<0.05 1C: Results for N1-MCL. CT>all,IT>EY,HE,CS,IS,IW,CL, IL,CA,IA, CW, CW > HE, IS, CL, IL, IA IW > HE, IS, CL, IL, IA, CS> HE, IS, CL, IL, CA, IA 1D: N1-MCM. CT>all, CW>EY,HE,CS,IS, IW,IT,CL,IL,CA,IA CS> HE, IS, IW, CL, IL, IA, IT>HE,IS, IL,IA, CA>IL 1E: N2-MC. CT>all, CW> EY,HE,CS,IS, IW,IT,CL,IL,CA,IA, IT>EY,HE,CS,IS,CL,IL,CA,IA, IW>EY,HE,CS,IS,CL,IL,CA,IA, CA>EY, HE, IS, IL 1F: N1-PPC. No significant differences other than all conditions from baseline. 1G: N2-PPC, CA> HE, IW, CT; CS> IW, CT, CL>CT, EY>CT. 1H-L: Event related averages for an example neuron from each brain area. Neurons selected are representative of the population. Colors show responses to different effectors. Mean and 95% confidence intervals are shown. H: N1-MCL, I: N1-MCM, J: N2-MC, K: N1-PPC, L: N2-PPC.

We asked the participants to attempt to move twelve effectors in five directions using a center-out paradigm where the effector to be moved was instructed on the screen (Supplemental Figure 1). For each movement, the direction was indicated by one of five targets changing color from gray to red. An inter-trial interval (ITI) followed each movement during which the participant was at rest. Array recordings were made simultaneously as the participants attempted movements of the eyes, head, contralateral/ipsilateral shoulder, wrist, thumb, leg, and ankle. Attempted movements for both participants result in overt movement for the eyes, head, and shoulders.

### Overlapping full body representations with differing population preferences in MC and PPC

Is there an effector-specific movement code implemented in MC or PPC? We examined the population responses for effector preferences. The strength of representation across the body was determined by finding the cross-validated distance between population firing rates during movements for each condition and firing rates during rest (during the ITI). Signals were spike sorted. In both participants, both MC and PPC showed population responses that were distinct from the baseline for all 12 effectors (Figure 1c-g, permutation test, p<0.05 for MC and PPC), however the relative strength of these representations across effectors were different between the two brain areas.

In N1-MCL, N1-MCM, and N2-MC, the contralateral thumb was significantly more strongly encoded than other effectors (aside from the contralateral wrist in N1-MCM and N2-MC, permutation test p<0.05) (Figure 1c-e). In N1-MCL the next strongest response was to the ipsilateral thumb and in N1-MCM it was to the contralateral wrist (Figure 1c and d). In N2-MC, the contralateral wrist, ipsilateral thumb and ipsilateral wrist all evoked responses stronger than any of the non-hand effectors (Figure 1e; permutation test p<0.05). In contrast, the population responses from both N1-PPC and N2-PPC generally did not show signficnat coding strength differences between effectors (exceptions noted in the figure caption, permutation test p>0.05) (Figure 1f-g).

To help qualitatively understand the population results, we visualized exemplar single-neuron firing rates grouped by the effector. In the single neuron examples for N1-MCL, N1-MCM, and N2-MC, the neurons respond strongly to the contralateral thumb, followed by the ipsilateral thumb and contralateral wrist (Figure 1h-j). In the single neuron examples for N1-PPC and N2-PPC, the neurons respond to one, two, several, and even all effectors. The chosen examples show a neuron in N1-PPC that is tuned across all twelve effectors, and one in N2-PPC that is tuned to half of the effectors (Figure 1k-l, more examples in Supplemental Figure 2).

### Single neurons respond to multiple effectors in both MC and PPC with distinct patterns of overlap

We next sought to better understand the neural architecture that supports coding for multiple body-parts in each area. Here we focus on two aspects of functional organization at the single neuron level. First, was there evidence for a systematic relationship between different effectors, e.g. if the preferred effector is the contralateral wrist, is it more likely that the neuron is tuned to the ipsilateral wrist? Second, if a neuron is tuned to multiple effectors, are the spatial tuning preferences similar?

We calculated the directional modulation strength of single neuron firing rates during movements of each effector. The strength was determined by subtracting a neuron’s firing rate in its worst direction from the firing rate in its best direction. Only neurons that were significantly directionally tuned to at least one effector were included in the results (shuffle test, p <0.05). First, we found each neuron’s most preferred effector, or the effector that resulted in the strongest modulation of that neuron’s firing rate (Figure 2a-e).

**Figure 2.**
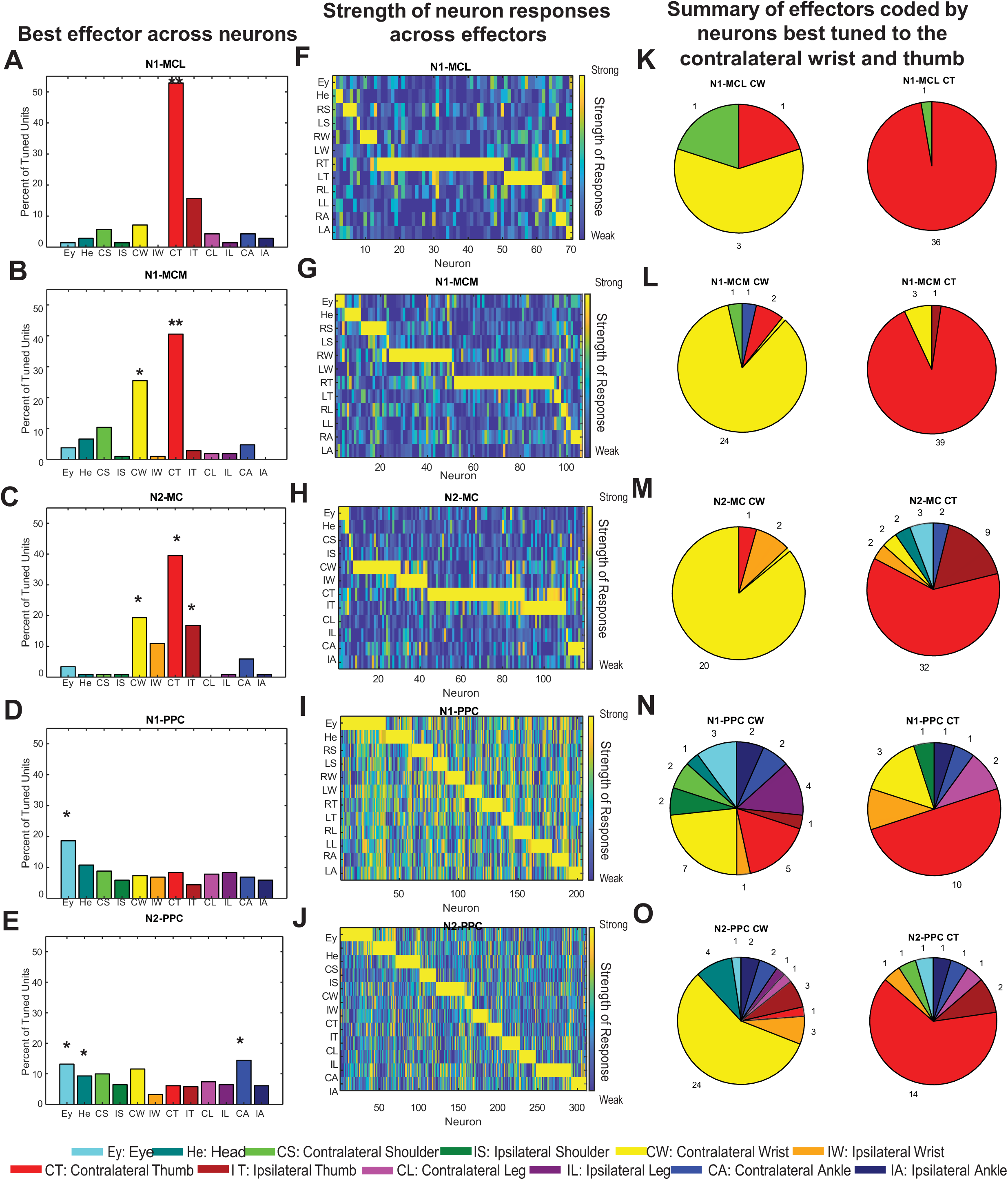
Single neuron responses show distinct patterns of overlapping representations across effectors in MC and PPC 2A-O: The strength of single neuron responses across effectors was found by determining each neuron’s best tuned direction and worst tuned direction, and then subtracting the firing rate during movements in the worst from the best for each condition. A-E: The best effector for each neuron was the effector that evoked the strongest response, or had the greatest difference in firing rates. Conditions are along the x-axis and the y-axis is the percent of neurons. A one-way anova was used to test for significant differences between the number of neurons best tuned to each effector. Two stars indicate that there were significantly more neurons best tuned to that effector than all other effectors. One star indicates that there were significantly more neurons best tuned to that effector than neurons best tuned to specific other effectors listed below. p<0.05.A: N1-MCL: CT>all. B: N1-MCM: CT> all, CW> all but CT. C: N2-MC: CT> EY,HE,CS,IS,IW,CL,IL,CA,IA; CW>EY,HE,CS,IS,CL,IL,IA; IT> EY,HE,CS,IS,CL,IL,IA. D: N1-PPC: EY> all but HE. E: N2-PPC: EY> IS,IW,IT,CT; H> IW,CT; CA> IS,IW,CT,IT,IL. F-J: The strength of a neuron’s response, with neurons sorted by their best effector and then by the number of effectors they coded. The strength of neural responses were normalized to the strongest response from each array. Each column shows a single neuron’s response across all 12 effectors. K-O: Breakdown of effectors coded by neurons best tuned to the contralateral wrist (left) and the contralateral thumb (right).

In each of the three MC arrays, around 40% of units preferred the contralateral thumb, in line with the population results (Figure 2a-c, N1-MCL: 53%, N1-MCM: 40%,N2-MC: 38%).

In the PPC arrays, neural preferences were spread across effectors, with between 4 and 18% of neurons best tuned to each effector (Figure 2 d, e). In both arrays the eyes stand out as being slightly more preferred than other effectors, and likewise for the contralateral ankle in N2-PPC, however this is only a significant difference for a few of the conditions (one-way ANOVA, p<0.05).

Next, we examined how the strengths computed for each neuron compared across all twelve effectors. The normalized strength of each neuron’s response is plotted with neurons sorted by two characteristics: their most preferred effector (from eyes to ankle); and the number of other effectors tuned (shuffle test, p<0.05; Figure 2f-j). The qualitative difference for MC and PPC is notable. To better understand the tuning of a single neuron across different effectors, we separated the neurons by their most preferred effector and then summarized what other effectors those neurons were tuned to in pie charts (Figure 2k-o; supplemental Figure 3). As shown in Figure 2d and e, there are a similar number of neurons that prefer each effector in PPC, but in MC there are very few neurons that prefer an effector other than the contralateral thumb or wrist (n<10). Thus, to compare single neuron tuning across effectors using a similar number of neurons recorded from arrays in both MC and PPC, the pie charts for neurons that most preferred the contralateral wrist and thumb are shown (Figure 2k-o, pie charts for other effectors Supplemental Figure 3).

In all three arrays in MC most neurons are tuned to one effector, and a smaller portion of single neurons are tuned to multiple effectors (Figure 2f-h & Supplemental Figure 4a-c). In the N1-MCL and N1-MCM arrays, less than 10% of neurons are tuned to multiple effectors (Supplemental Figure 4a & b). In N1-MCL only a single neuron that preferred the contralateral thumb was tuned to another effector (contralateral shoulder) (Figure 2k). There were only 5 total neurons that preferred the contralateral wrist in N1-MCL, with one of these neurons also coding for the contralateral shoulder, and another for the contralateral thumb. In N1-MCM, less than 10% of neurons that prefer the contralateral thumb were tuned to additional effectors, and this included the ipsilateral thumb and contralateral wrist (Figure 2l). In N1-MCM only four neurons that preferred the contralateral wrist were tuned to other effectors, and two of these were tuned to the contralateral thumb (Figure 2l).

In N2-MC, the most apparent effector overlap for single neurons is between corresponding ipsilateral and contralateral effectors, in particular the wrist and thumb (Figure 2 h, m). Around 40% of the neurons that were best tuned to the contralateral thumb were tuned to additional effectors. Of these, just under 50% are tuned to the ipsilateral thumb. Two of the three neurons that prefer the contralateral wrist and are tuned to multiple effectors are tuned to the ipsilateral wrist, with the third neuron tuned to the contralateral thumb (Figure 2 m). Overall, across all MC arrays, tuning across multiple effectors by a single neuron was largely restricted within a single limb with few exceptions.

In both N1-PPC and N2-PPC, a large portion of neurons are tuned to multiple effectors (Figure 2i, j and Supplemental Figure 4 d & e-49% in N1-PPC, 35% in N2-PPC). Additionally, there is no clear consistent relationship between the effectors to which a single neuron is tuned (Figure 2i-j). Rather, neurons show clear preferences for what appear to be random sets of effectors. In both N1-PPC and N2-PPC, neurons that preferred either the contralateral wrist or contralateral thumb were tuned to the full range effectors from head to ankle (Figure 2n-o, Supplemental Figure 3 shows same pattern across all effectors).

Comparing the strength of single neuron responses across all effectors in each brain area, we found a trend toward stronger representation across all effectors in PPC than MC (Supplemental Figure 5). To ensure these effects were truly driven by single neurons, we applied a stricter criterion for spike quality to rule out any potential signals from multi-unit activity and found the same pattern of results (Supplemental Figure 6 a-e).

Next, we compared the directional tuning properties of neurons across effectors. We asked whether single neurons used the same spatial code for different effectors. We found the linear regression coefficients for each neuron and used those coefficients to determine the angle between the directions coded in the response for each effector. An angle of zero implies that the neuron used the same directional tuning across effectors. This comparison was done for the most and second most preferred effector for each neuron (determined using the criteria from Figure 2a-e). Few neurons from N1-MCL and N1-MCM were tuned to more than one effector meaning no strong claims can be made about the directional tuning of these neurons (Figure 3a & b, n=4 for N1-MCL and n=12 for N1-MCM). In N2-MC, N1-PPC, and N2-PPC, we found most neurons are tuned to the same direction across effectors, meaning zero angle of difference (Rayleigh test for uniformity p<0.05; circular mean N2-MC = 0.000° ± 18.5°, N1-PPC = - 0.493° ± 4.412°, N2-PPC = 0.000° ± 4.84°; Figure 3c-e).

**Figure 3:**
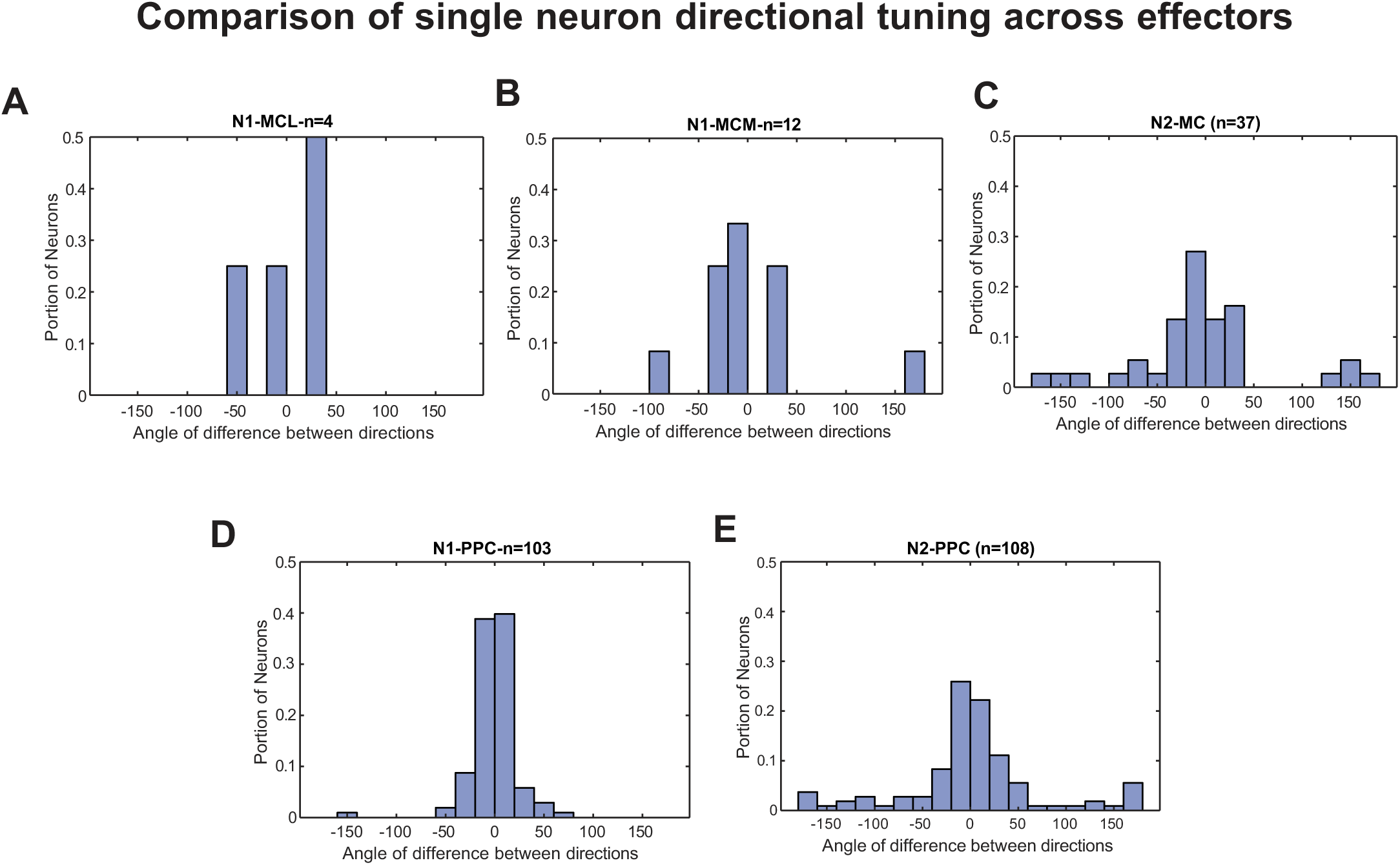
Preserved directional tuning across effectors within single neurons A–E The angle between the direction vectors coded for the most preferred and second most preferred effector of single neurons. A. N1-MCL circular mean (cm) = 0.344°, confidence intervals (ci) = 48.129°. Rayleigh test for uniformity p < 0.05. B. N1-MCM cm = -11.05°, ci = 31.341°. Rayleigh test for uniformity p < 0.05. C. N2-MC cm = 0.000°, ci = 18.5°. Rayleigh test for uniformity p < 0.05. D. N1-PPC cm = -0.493°, ci = 4.412°. Rayleigh test for uniformity p < 0.05. E. N2-PPC cm = 0.000°, ci = 4.84°. Rayleigh test for uniformity p < 0.05.

Finally, we asked what reference frame neurons used to encode direction. Spatial information can be represented intrinsically, relative to the location of our body and joints, or extrinsically, relative to location in the external world. Here we leveraged the fact extrinsic versus intrinsic coordinates are naturally dissociated between the left and right sides of the body (see Supplemental Figure 7a, b). We compared the linear regression coefficients found above for ipsilateral and contralateral pairs of effectors to determine the reference frame used by each neuron. Again, in N1-MCL and N1-MCM there were too few neurons to determine the reference frame this way (n<=2). In N2-MC, there was a significantnegative correlation between the coefficients for pairs of contralateral and ipsilateral effectors, demonstrating an intrinsic reference frame (Supplemental Figure 7c, Pearson correlation=-0.71, p <0.001). In contrast, N1-PPC shows a positive correlation between contralateral and ipsilateral coefficients, suggesting an extrinsic reference frame (Supplemental Figure 7d, Pearson correlation, r=0.94, p <0.001). Finally, in N2-PPC there was no trend toward a positive or negative relationship, implying a mixture of both intrinsic and extrinsic reference frames (Supplemental Figure 7e, Pearson correlation, r=-0.10, p=0.6).

### Arbitrary vs specific shared information

How might MC and PPC be using these overlapping effector representations? One possible explanation discussed in motor literature is that this provides a shared architecture through which learned motor skills can be transferred across limbs ^9, 22, 26^. At the level of effector preference, we report clear distinctions between representations in MC and PPC. While these differences in the structure and extent of overlap in effector representation give some intuition, they do not answer the question of how efficiently information can be shared across effectors in either area. We call the extent to which information is shared between effectors the transfer efficiency. If the neural code has high transfer efficiency across effectors, information is shared between two effectors regardless of their location on the body or function in movement. Another possibility is that high transfer efficiency only exists between specific effectors, for example effectors that are more likely to be moved together such as the contralateral wrist and the contralateral thumb, or the contralateral and ipsilateral wrist for bimanual tasks.

To assess how information was shared across effectors in MC and PPC, we trained a partial least squares linear regression model to find a directional coding subspace that was shared for two effectors and not the rest. In other words, we looked for a subspace where the spatial code was approximately the same for two effectors, but distinct from other effectors. This was done for the 66 possible pairings of effectors. To evaluate the transfer efficiency of information between these effector pairs, we found the correlation matrix for actual vs model-predicted position. Pairs with high transfer efficiency will have high R^2^ values, and the average R^2^ across pairs gives a sense of the population’s overall transfer efficiency. To determine significance, this R^2^ value was compared to a null distribution of R^2^ values that was generated for each pair by shuffling the firing rates for trials with one effector, to mimic a population where no information is shared between effectors. Finally, to be sure this pattern reflected sharing between individual neurons, we excluded signals that may be the result of multiple neurons rather than isolated single neurons.

We found that only specific pairs of effectors significantly share directional coding in MC (Figure 4a-c). In N1-MCL and N1-MCM the highest transfer efficiency between two effectors was between the contralateral thumb and wrist, and in N2-MC for the contralateral and ipsilateral thumb. The fewest pairs were found to share information in N1-MCL, with only three effector pairs sharing spatial information. In N1-MCM, the contralateral thumb shares information with most contralateral effectors and most contralateral effectors shared with each other (with a few exceptions). In N2-MC the contralateral wrist, thumb, and ankle shared information with each other, and with their ipsilateral counterparts. The shared information between the thumb and other hand effectors was somewhat expected based on the other results seen in MC thus far, but finding the ankle also shared with other effectors was more surprising.

**Figure 4:**
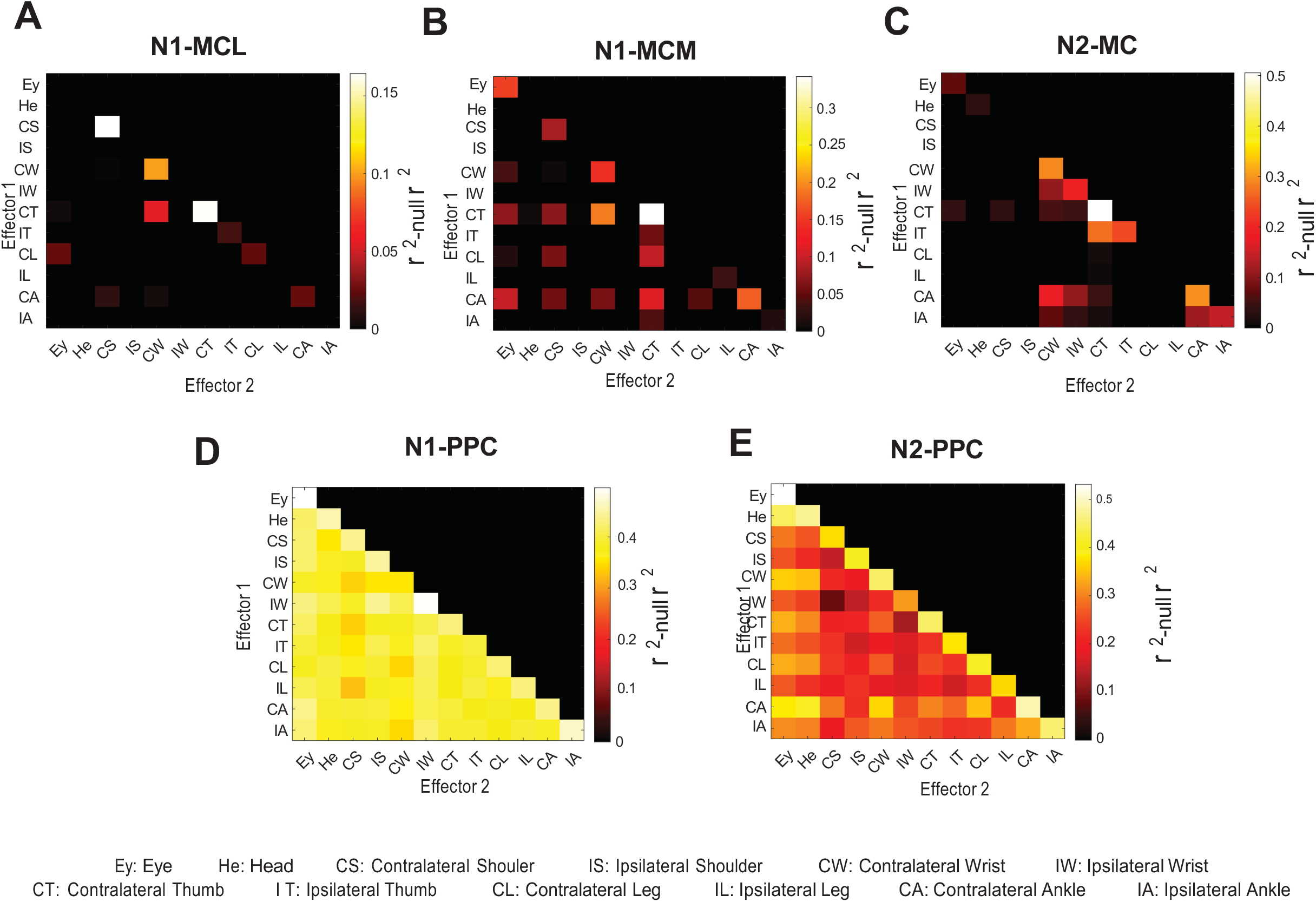
Evaluating shared information between effectors A PLSR model was trained using firing rates from all neurons, and position information for two effectors, for each possible pairing of effectors. The positions predicted by the model were then correlated with the original input position values to determine whether spatial information had been shared between the two effectors. This same process was done for a matrix of shuffled firing rates to obtain a null distribution to compare these model’s performances with for significance. A-E: Performance of models trained for each effector pair. The x- and y-axes are the two effectors used in the pair. The diagonal is made up of a single effector rather than a pair of effectors. The threshold for significance determined by the null r2 distribution is marked on the colorbar. Only the lower triangle is shown because the matrices are symmetric. A: N1-MCL, B: N1-MCM, C: N2-MC, D: N1-PPC, E: N2-PPC.

Previous work has shown that certain motor skills like writing can be transferred from the hands to the feet. Perhaps this shared spatial information reflects the functional organization that has also been reported alongside somatotopy in MC ^8^. The interpretation of this finding will be expanded on further in the discussion.

In N1-PPC and N2-PPC, shared directional coding is found for every set of effectors (Figure 4e-f). All effector pairs seem to share information relatively equally especially in N1-PPC. Although significant sharing is found for all pairs in N2-PPC as well, more variation is noticeable across pairs. This could be a result of the mixed intrinsic and extrinsic reference frames used, which were not fully accounted for in this analysis.

## Discussion

We found that the entire body is represented within small patches of hand areas in human MC and PPC. These results add to the growing body of work that illustrate a more complex organization of motor areas than initially expected ^6, 7, 9, 18, 22^. Importantly, we found clear differences in the structure of these representations in each area that ultimately align with previous notions of distinct functional organization within low and high level motor areas.

In the lower level MC, we found a clear dominant effector, the contralateral thumb. In addition, the pattern of overlapping effector representations seen also fits with a broader somatotopic organization. For example, the prominent population response to the contralateral wrist supports a gradual transition between coding for neighboring parts of the body. Further, neurons tuned to the contralateral thumb were more likely to be tuned to the contralateral wrist than most other effectors. Another pattern that emerged in both population and single neuron preferences was overlapping representation of the contralateral and ipsilateral sides for both the wrist and thumb. Although this strays somewhat from a classic somatotopic view, coding for both sides of the body has also been reported in NHPs ^11–14^. More work on bimanual coordination is needed to further understand the role of ipsilateral hand representations in both NHPs and humans. Ultimately, our results suggest that despite some complexity at the level of single-neuron responses, MC still follows a broadly somatotopic organization where different parts of the body are emphasized in different cortical locations. These findings do not rule out the possibility of other parts of MC demonstrating more functional organization, as fMRI work suggests that some locations within MC show more anatomically specialized representations than others^6^.

In higher level PPC, we observed an equal strength in representation across effectors, although not to the extent of an effector agnostic representation. Initial work in PPC of NHPs established separate anatomical patches for coding eye, hand, and arm movements; however, in more recent fMRI work in humans, such effector-specific regions have remained elusive ^27–29^. Further, studies that compare hand and foot movements report areas in PPC that are activated equivalently by both movements ^18, 21^. Overlap between effectors has also been found in single neurons, where previous recordings in human PPC from our lab found the contralateral and ipsilateral hand and shoulder encoded in the same, small patch of cortex ^22^.

The present findings tie these results together and reveal an important insight about the initial interpretation from fMRI studies. Instead of a purely effector or functional organization within PPC, when recording from the single neuron level, we found that individual effectors are randomly mixed. This ultimately results in an equal strength of coding across effectors within the population, which appears as an effector-agnostic population code when observed with the spatial resolution of fMRI.

What is the functional purpose of the overlapping effector representations seen in each area? In motor literature, it has been suggested that this could facilitate the transfer of motor skills from one limb to another ^9, 22^. Transfer of motor skills can occur between limbs in motor-intact individuals, and not only are skills transferable, but often, an individual’s movement characteristics are preserved from one body part to the next ^30^. An example of this is in handwriting, where it has been found that writing produced by the digits, wrist, arm, and foot all share common characteristics ^31^. Further, both amputees and individuals born without hands show great dexterity with the feet when used in place of the hands ^26^. This suggests that at least some motor brain areas could contain a shared substrate for multiple effectors. PPC has been identified as an area of particular importance for such a process from fMRI evidence both in amputees and motor intact individuals ^21^. For example, one fMRI study comparing finger and foot movements during name signing and zig-zag motions found increased activation of SPL regardless of effector or condition ^32^. Our results sharpen this prior work, demonstrating that what is shared is not simply a common spatial encoding, but, rather, a more specific relationship between distinct sets of effectors.

Another potential role for the coding structure is computational efficiency. PPC has been shown to implement a coding strategy called partially mixed selectivity, where some variables are coded randomly (or nonlinearly) with respect to each other across the population, while other variables are coded with some structure ^22, 33–35^. For example, in our results we find that single neurons code for random combinations of effectors; however, a single neuron tends to use the same spatial/direction code across the effectors it encodes. Mixed selectivity in other cortical areas like prefrontal cortex has been suggested to provide a more efficient way of representing more complex and/or higher cognitive information, which would suggest PPC having a higher-level representation of motor control ^35, 36^. This notion is further supported by results from fMRI studies that have found similar activity in PPC during overt movements to that during observed and imagined movements, but not as much so in MC ^37–43^. Given that PPC is an area with great variety in the types of information it represents from motor control to semantics, there is still much to be explored when it comes to understanding its functions ^44–46^.

Another possible function of overlapping representations is coordinating movements of multiple effectors. It has been proposed that during hand-eye coordination, information about the direction of both hand and eye movements are combined through global tuning fields, or the shared directional coding of individual cells in PPC for the hand and eyes ^47^. While in this study we do not explicitly test coordinated movements, the common spatial tuning used by single neurons across effectors appears to extend the concept of global tuning fields to humans and to effectors across the entire body.

One question still left open is whether our findings for attempted movements in individuals with paralysis extend to overt movements. Evidence from fMRI work in motor intact individuals largely supports this to be the case ^6, 8, 18, 21^. Also of note is the potential impact of precise implant location on effector specificity. Indeed, while the same brain regions were targeted for both participants, there were slight differences between the patterns of overlap observed. Further, in both MC and PPC, much work emphasizing effector separation has been within the sulcus (central and intraparietal respectively). Our recordings are on the surface. That said, both fMRI and sEEG have recently revealed highly overlapping regions within the central sulcus as well ^6, 7^. Additionally, identifying effector specific regions in human PPC using fMRI work in human has revealed more flexibility than specificity ^27, 29^.

This study presents the first single neuron evidence for encoding of effectors across the body in human posterior parietal cortex. In addition, we recorded from motor cortex and posterior parietal cortex simultaneously, allowing us to compare the encoding of movement variables in distinct neural populations. The surprising nature of these results contributes to the growing body of work that suggest that designing simplified and highly controlled experiments to probe predefined functions determined by existing cortical maps may limit our understanding of the more complex coding properties of different areas of cortex ^48, 49^. These results also have implications for evaluating the use of different brain areas in the control of therapeutic devices with brain-machine interfaces for those with impaired movement. In MC, strong representation of single effectors supports its usefulness in biomimetic control. In PPC, equal representation across effectors highlights its potential to be used flexibly across several applications.

## Methods

### Study participants

The present data was collected from two participants enrolled in a BMI clinical study (ClinicalTrials.gov Identifier: NCT01958086). The institutional review boards of California Institute of Technology, Casa Colina Hospital and Centers for Healthcare, and University of California, Los Angeles approved study procedures including informed consent, implant surgery, and experiment design. Participant N1 is a right-handed male with a C3-C4 level spinal cord injury that occurred approximately ten years before his enrollment in the study. N1 can move his eyes, head, and shoulders (he can move his shoulder but cannot stabilize the arm). In addition, N1 shows weak residual movements (twitches) of the wrist and thumbs.

Participant N2 is a right-handed male, with a C4-C5 level spinal cord injury that occurred approximately three years before his enrollment in the study. N2 can move his eyes, head, and shoulders like N2.

### Experimental Procedures

Two adaptations of a traditional center-out task were used. Both tasks were displayed on a screen in front of the participant. Participants were instructed to attempt movements as though they were motor-intact. At the start of each trial, participants were asked to have the effector that was going to be moved in a neutral position and their eyes fixed in the center of the screen. In the first task, the participants were instructed to attempt movements of twelve different effectors (head, eyes, right/left shoulders, wrist, thumbs, legs, and ankles) in five different directions, 72 degrees apart. The direction of the movement was instructed by one of five targets on the screen changing color from gray to red as a “go” cue (Supplemental Figure 1a). The participants were asked to make the instructed movement and then relax. They moved one effector to each of the five targets before the instructed effector changed to the next. Fixation was kept at the center except during trials for the eyes. Between each trial was an inter-trial-interval (ITI) during which time the participant was at rest. Each ITI was 1.5s and each go phase was 2s. A total of 7 sessions were collected for N2 and 6 for N1. Three runs of the task with two repetitions for each movement were collected for each session.

### Implant Locations

Participant N1 was implanted with four 64-channel NeuroPort Utah electrode arrays in his left hemisphere (fig 1a). Two arrays were placed near the hand knob of left precentral gyrus, with one slightly more lateral and the other slightly more medial (denoted N1-MCL and N1-MCM respectively). A third array was placed in the superior parietal lobule (N1-PPC). A final fourth array was placed in the supramarginal gyrus, however results from this array are excluded from the present study because only a limited number of tuned single neurons were recorded. Data from participant N1 was collected ranging from three to six months after implant surgery. Participant N2 was implanted with two 96-channel NeuroPort Utah electrode arrays in his left hemisphere (fig 1b). One array was placed near the hand knob of the left precentral gyrus and is referred to as motor cortex or “N2-MC”. The second array was implanted in the left superior parietal lobule and is denoted “N2-PPC”. Data from N2 was largely collected 20-22 months after surgery (along with one session 13 months and a final session 27 months post implant). A presurgical functional MRI during which each participant performed a grasping imagery task showed substantial bold response in regions of each implant location.

### Neural signal recording and preprocessing

Microelectrode arrays in each participant were recorded simultaneously. Neural signals were amplified, bandpass filtered (0.3 Hz–7.5 kHz) and digitized (30 kHz, 16 bits/sample) (NeuroPort Neural Signal Processors, Blackrock Microsystems Inc.). To detect action potentials, the signal was then high-pass filtered at 250 Hz and thresholded at -3.5 times the root-mean square voltage (for each electrode). We used the k-medoids clustering method along with the gap criteria in order to identify single neurons and the total number of waveforms respectively ^50^. This was done for the first four principal components (selected to account for 95% of waveform variance).

For analysis of each behavioral epoch during the 12-effector task, we calculated firing rates from neural activity over a 500ms window of neural activity. The movement execution (‘Go’ or ‘move’) analysis window was defined as the 500 ms window starting 250 ms after the Go cue. Similarly, the 500 ms ITI window used in the distance analysis began 250 ms into the ITI phase.

Neurons with an average firing rate of less than 1 Hz across the entire task were excluded from the analysis as noise. Additionally, all analysis was repeated on well-isolated neurons to avoid influences from multi-unit clustering. We classified neurons that corresponded to the lowest third of L-ratio values as well-isolated^51^.

### Cross-validated distance analysis for population and single unit firing rates

The cross-validated mahalanobis distance^52^ was used to compare the population level neural activity during movements of each effector and a baseline (no movement). Leave-one-out cross validation was used. A permutation test with 1000 shuffles was used to determine whether there was a significant difference between the distances of firing rates from baseline during movements of each effector.

### Event Related Averages

Firing rates for each neuron were grouped by condition and then the mean and standard error of the mean of these responses were found for 100ms bins across a 2.5s window.

### Strength of single unit responses across effectors

To determine the strength of an individual neuron’s response to different effectors, first we determined a single neuron’s cross validated best and worst tuned direction by finding maximum and minimum firing rates across trials. The response for each effector was then evaluated by subtracting the neuron’s firing rate during its best tuned direction from the neurons firing rate during its worst tuned direction during movements of each effector. To evaluate whether the response to an effector was significant, the same measure was performed with shuffled firing rates to find a null distribution for that neuron to compare the strength of response for each effector (>95 percentile). The best effector for each neuron was then determined by finding the largest difference in firing rates between best and worst direction. Main figure results were also normalized and neural responses were sorted first by best effector and then the number of effectors to which that neuron was tuned (least to most).

### Comparing spatial tuning of single neurons across effectors

The spatial tuning of each neuron was determined independently for the most preferred and second most preferred effector for each significantly tuned neuron (most preferred, second most preferred, and test for significance described in the above section). This was done by performing a linear regression using neural firing rates as independent variables and x and y values for the target location on the screen as response variables. The coefficients from this regression give the directional vector coded by each neuron. The angle between these resulting vectors was calculated to compare the spatial tuning between a neuron’s most preferred and second most preferred effectors. Finally, the MATLAB toolbox CircStat was used to calculate the circular mean and confidence intervals for each distribution, and to perform the Rayleigh test for uniformity.

### Determining body vs world-centered coordinates

To determine whether neurons were encoding space in a body-centered, or intrinsic reference frame, or a world-centered, or extrinsic reference frame, linear regression was used to find the direction vector coded by each neuron for pairs of contralateral and ipsilateral effectors (described in the section above). Then the correlation coefficient values from these regressions were correlated for each pair of effectors. A positive correlation indicated extrinsic coding, a negative correlation indicated intrinsic coding and no correlation implies some combination of the two. We found a significant negative correlation in N2-MC that implies intrinsic coding of direction (supplemental figure 7). Thus, in any of the analyses where we want to compare between directional coding of the contralateral and ipsilateral sides of the body, we must adjust the position information to be the same in intrinsic rather than extrinsic coordinates (Figures 3,4). This can be achieved by switching the position information for movements to the left and right targets.

This was only done for N2-MC, because no such intrinsic relationship was found in the other arrays.

### Finding shared subspaces between effectors within the population code

We evaluated the structure of the neural population code using a cross-validated partial least squares regression model to find shared directional subspaces between pairs of effectors, for each possible combination of effectors (Figure 4). A series of models for each effector pair were trained using neural firing rates across all units as predictor variables and the target position information for the two effectors as the response variable for a subset of the trials. To control for effects of training dataset size, these subsets were created by randomly choosing fifteen trials for each of the effectors in the pair but using all thirty trials for the models trained on a single effector. The model output was the predicted position information. The performance of each model was evaluated by taking the bootstrapped correlation between the input labels and those predicted by the model. The mean value of the correlation of the models of 200 randomized subsets was calculated for each effector pair. To determine whether the r2 values were driven by shared information or a single effector, a null r2 distribution was generated. The null r2 distribution was found using the same general steps, but with randomly shuffled firing rates for trials for one of the effectors for pairs of effectors, or half of the trials for the single effector cases. This was done to mimic an effector pair where one effector may have spatial information, but that information is not shared between the two effectors. The threshold for significance for shared information was p<0.01. Each effector pair generated two r2 values (for each order of pairing), and to be considered significant, we strictly required p<0.01 for both orderings. In addition, effector pairs where one or both effectors did not significantly share information within itself were excluded for their lack of spatial information. Both models were k-fold cross-validated over 6 folds. To account for the intrinsic reference frame in N2-MC, the position information for right and left targets was switched for ipsilateral effectors.

## Data Availability

All data produced in the present study are available upon reasonable request to the authors

## Acknowledgements

We would like to express our gratitude to participants N1 and N2 for making this research possible. We also thank Viktor Scherbatyuk and Evangelia Tsolaki for technical assistance. This work was supported by National Eye Institute grant UG1EY032039, Tianqiao and Chrissy Chen Brain-Machine Interface Center at Caltech, Swartz Foundation, and Boswell Foundation.

