## Supplemental Figures for "Distinct patterns of whole-body coding in human motor cortex and posterior parietal cortex"

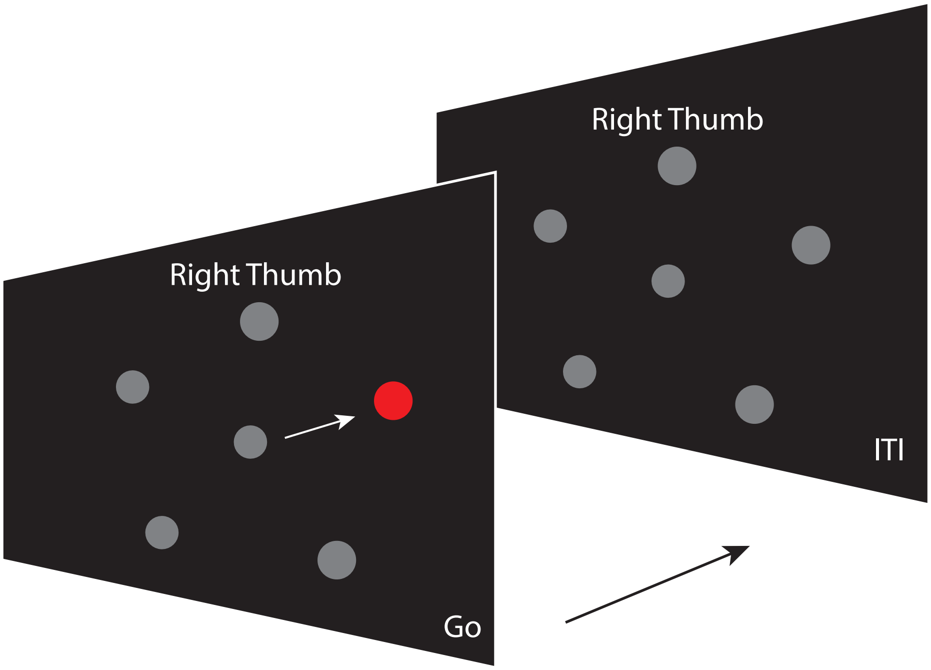


Supplemental Figure 1
Example frames from the twelve effector center-out task. In this task, participants moved one of twelve possible body parts in five directions (shown by the gray targets) before moving on to the next effector. The effector being moved was instructed by text on a screen in front of the participant, and motor initiation was cued when one of the five outer targets changed from gray to red. Movement execution phases were 2s and followed by 1.5s of rest during an inter-trial-interval.

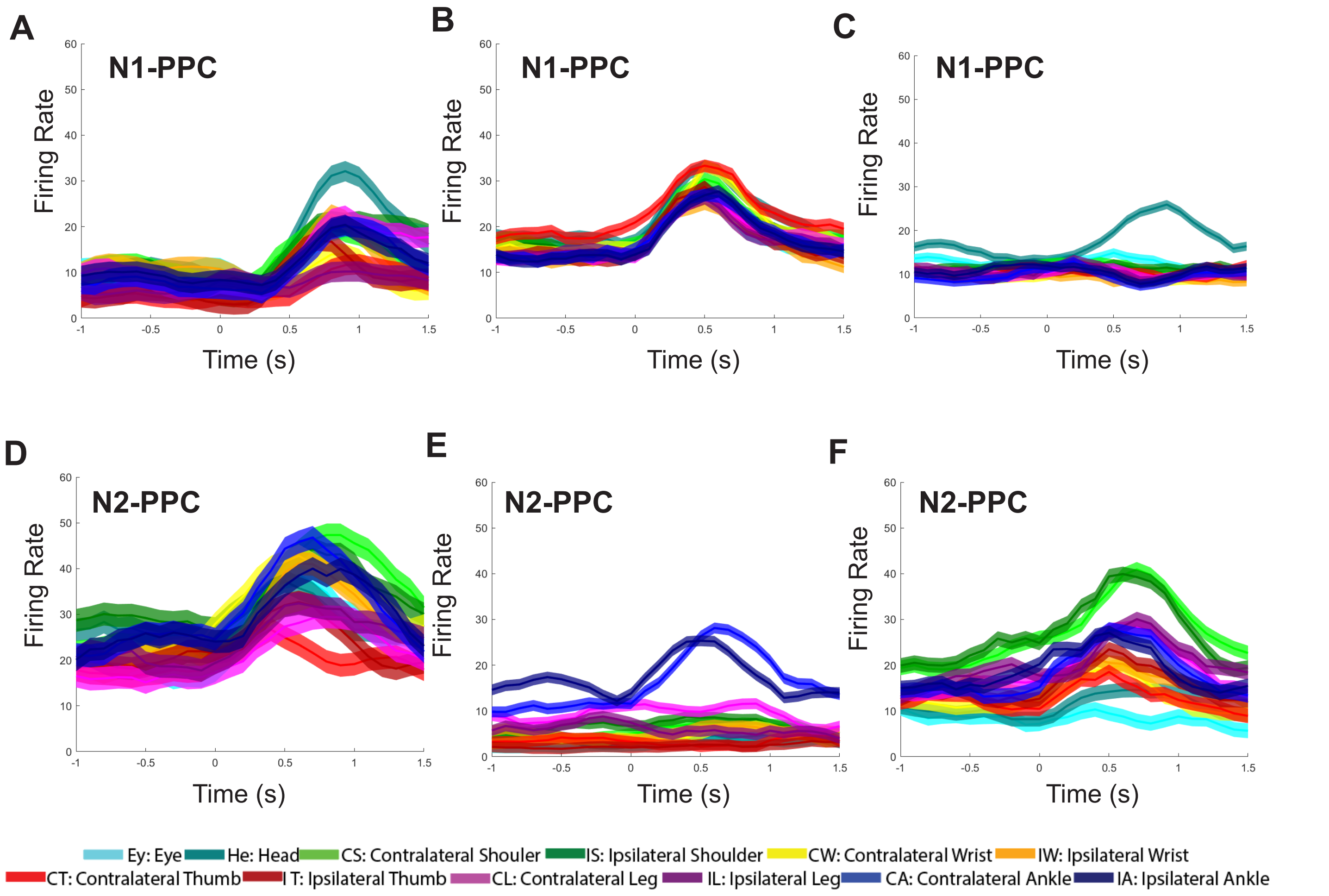


Supplemental Figure 2
Event- Related Averages for more example neurons. A-C examples from N1-PPC. D-F examples from N2-PPC.

### Summary of effectors coded by neurons in N1-PPC and N2-PPC separated by the best-tuned effector for each neuron

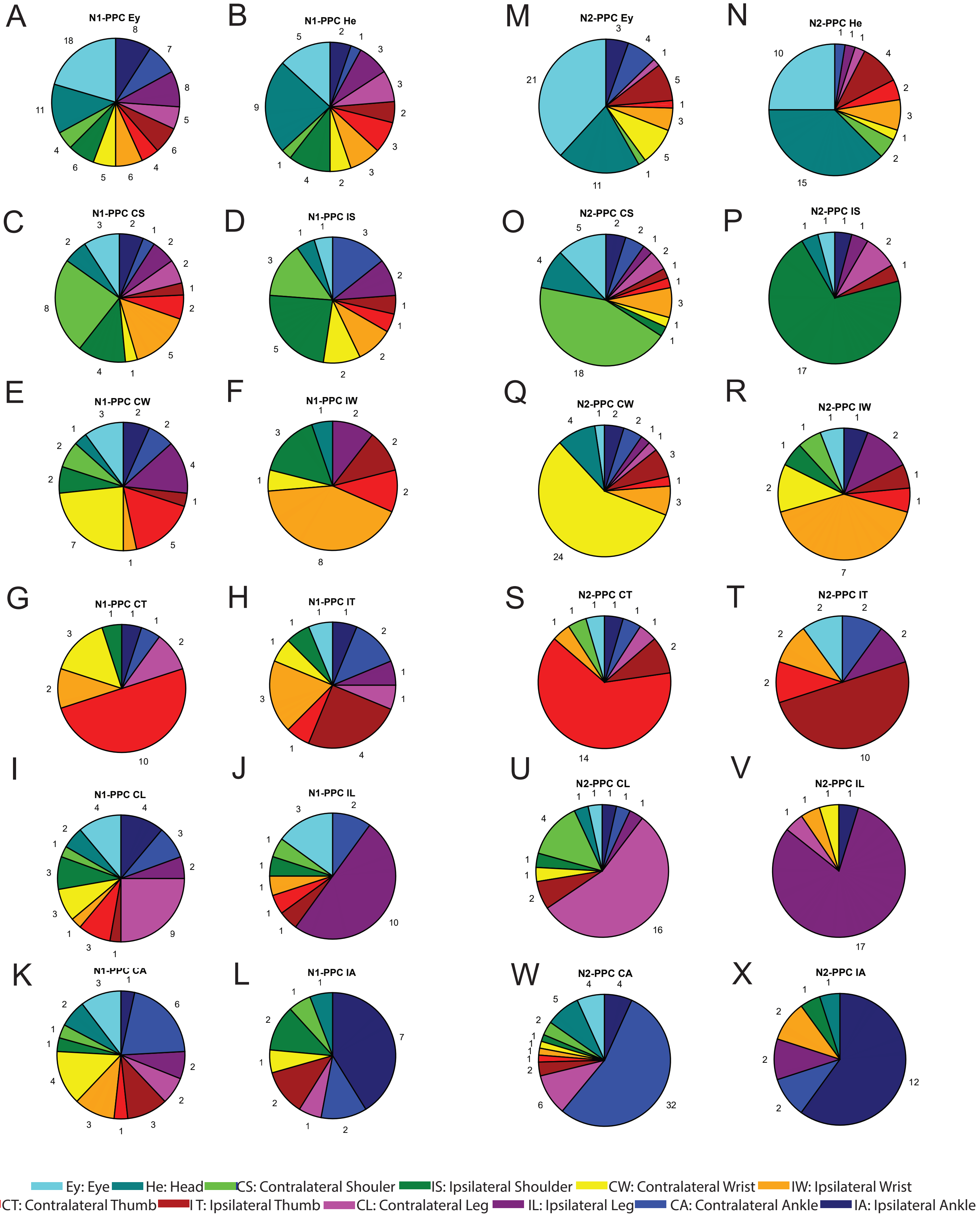


Supplemental Figure 3 Breakdown of single neuron tuning preferences for all effectors.
Neurons are sorted into groups based on the effector they most preferred. The different effectors coded by neurons with each effector preference are summarized in eaech pie chart. A-L: N1-PPC, K-X: N2-PPC.

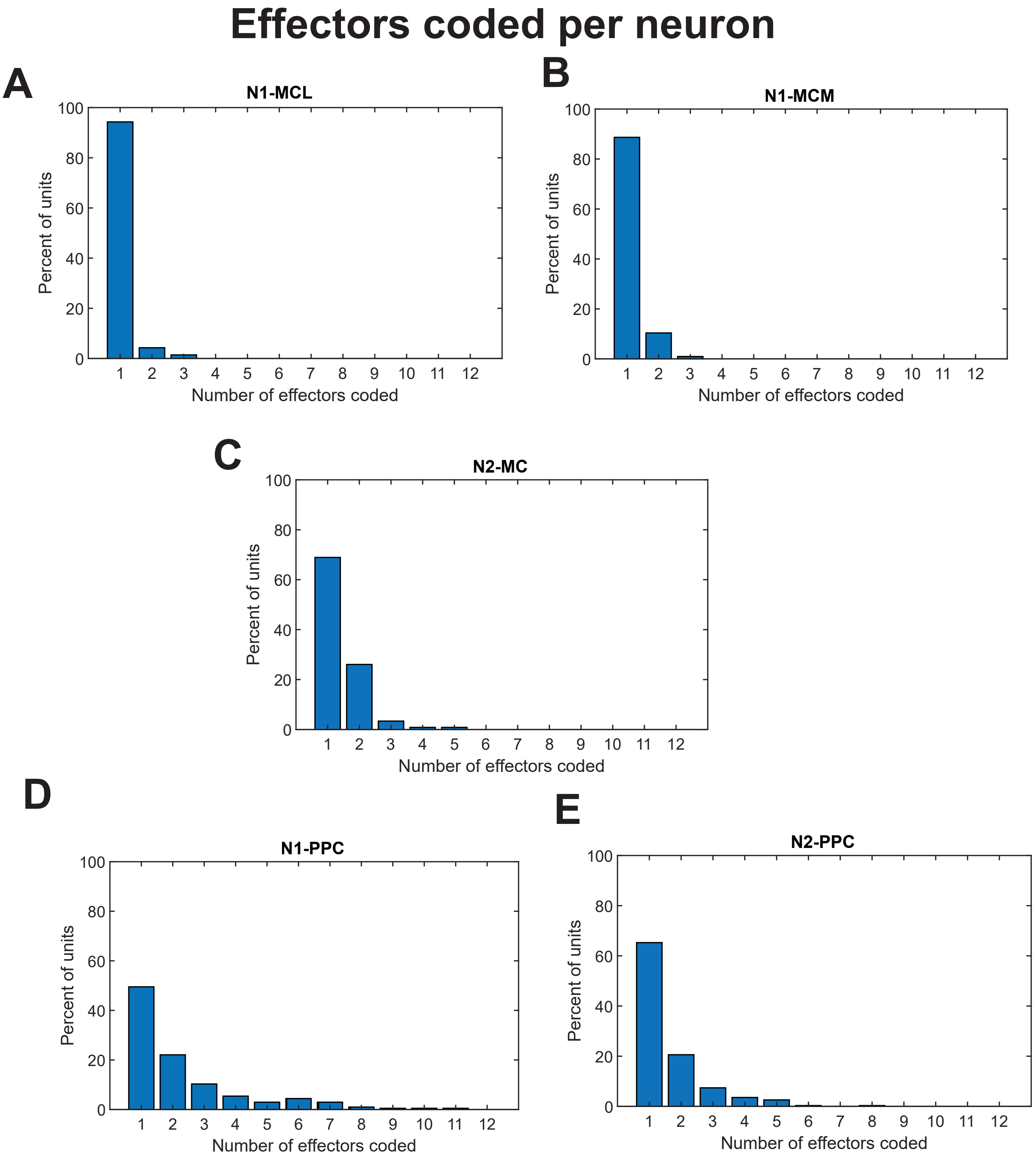


Supplemental Figure 4
The number of effectors coded by each neuron was found by determining the number of effectors that evoked a significantly strong response from that neuron (as computed for figure 2). A: N1-MCL, B: N1-MCM, C: N1-MC, D: N2-PPC,E: N2-PPC.

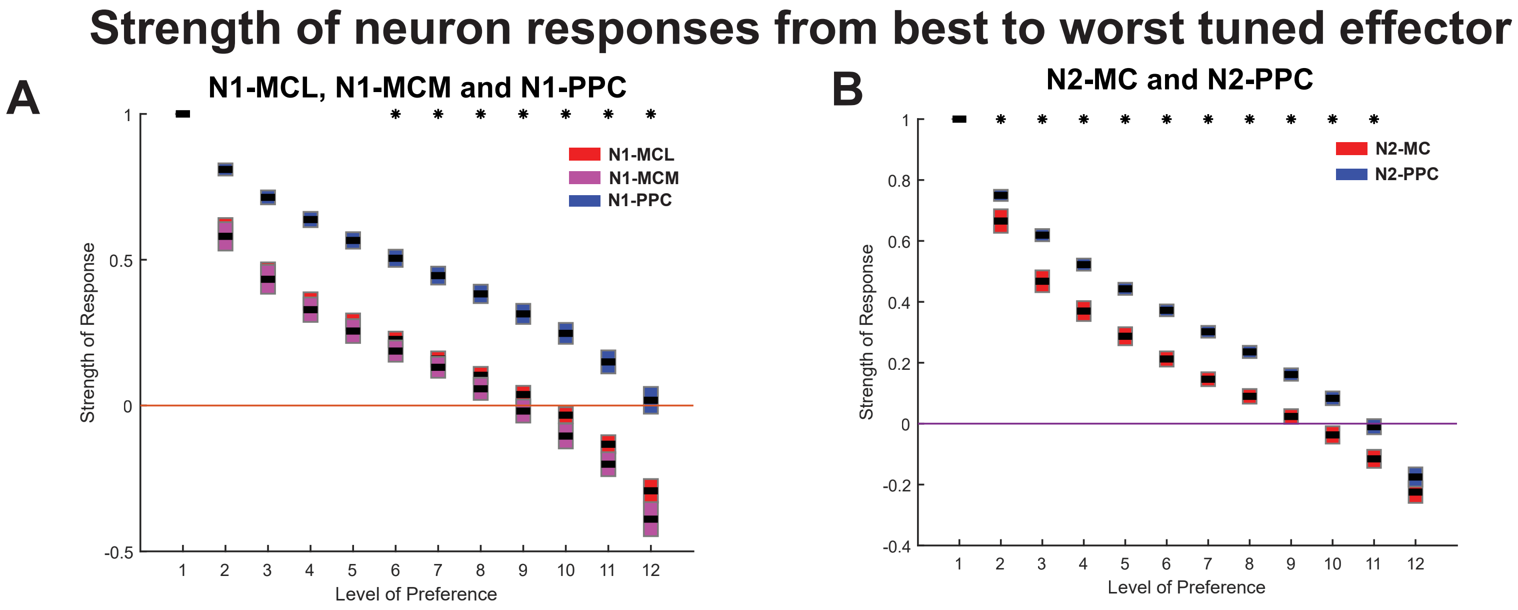


Supplemental Figure 5. Responses across effectors are stronger in PPC than MC
A & B The strength of each neuron's response across all twelve effectors was sorted from strongest to weakest response. Then the distribution of responses across the best effector, second best effector, and so on, was plotted for each array. A: Distribution of responses from N1-MCL (red), N1-MCM (magenta), and N1-PPC (blue). Stars indicate differences between N1-MCL and N1-PPC as well as N1-MCM and N1-PPC. B: Distribution of responses from N2-MC (red) and N2-PPC (blue). Stars indicate differences between N2-MC and N2-PPC.

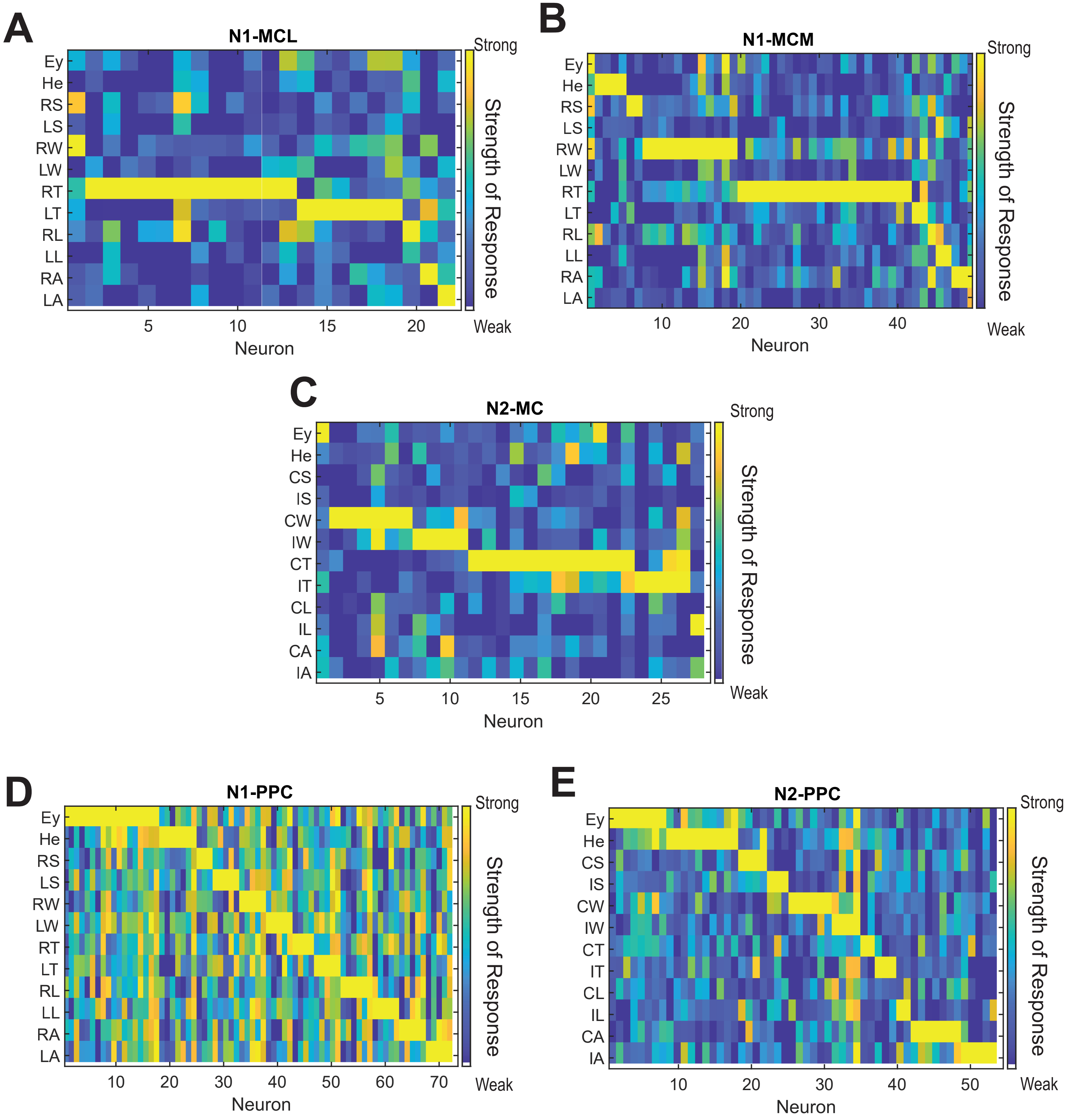


Ey: Eye He: Head CS: Contralateral Shouler IS: Ipsilateral Shoulder
CW: Contralateral Wrist IW: Ipsilateral Wrist CT: Contralateral Thumb I T: Ipsilateral Thumb
CL: Contralateral Leg IL: Ipsilateral Leg CA: Contralateral Ankle IA: Ipsilateral Ankle

Supplemental Figure 6 Responses from well isolated units
A-E: The same strength of response used in figure 2F-J was found for neurons that met a more rigorous criteria in spike quality. A: N1-MCL, B: N1-MCM, C: N2-MC, D: N1-PPC,E: N2-PPC.

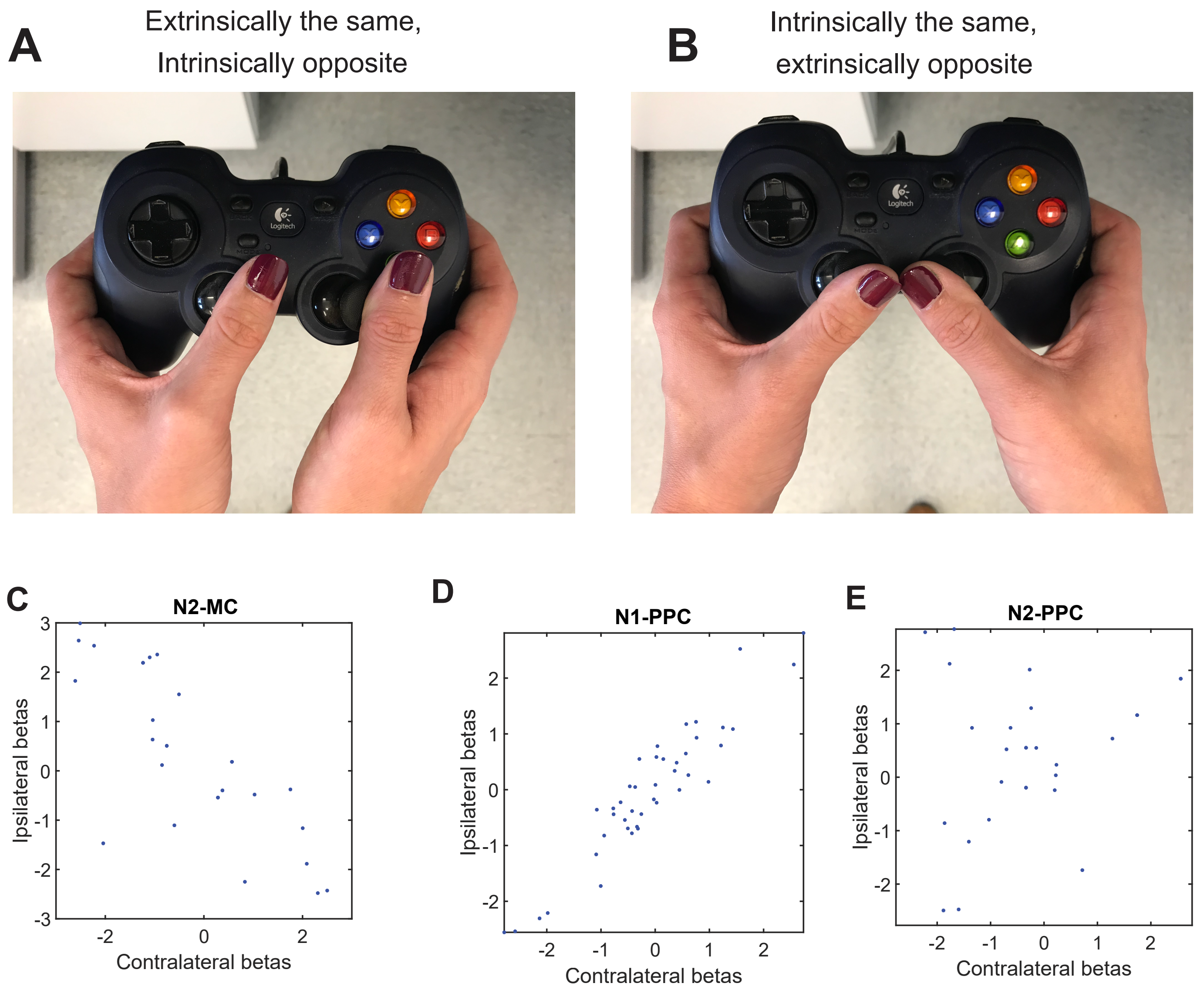


Supplemental Figure 7

A. An example of thumb movements that are both extrinsically in the same direction, to the right, but they require movements in opposite direction relative to the joint of the thumb, or intrinsically in opposite directions. B An example of movements that are in extrinsically opposite directions (ipsilateral thumb to the right, contralateral thumb to the left), they require movement in the same direction relative to the thumb joint, and are intrinsically the same. C – E The resulting betas for linear regressions for contralateral and ipsilateral pairs of effectors. C N2-MC $r^2 = -0.75$, $p < 0.001$. D N1-PPC $r^2 = 0.94$, $p < 0.001$. E N2-PPC shows no correlation.
